# Psychological influences on hypertension in Southeastern Nigeria: a mixed-methods study protocol

**DOI:** 10.64898/2026.02.15.26346343

**Authors:** Awele Rosemary Ebigwei Omeda, Marcus Chilaka, Masoud Mohammadnezhad, Eleftheria Vaportzis

**Author notes:** Corresponding author (EV). **Funding:** The authors received no specific funding for this work. **Competing interests:** The authors have declared that no competing interests exist. **Data Availability:** No datasets have yet been generated from this protocol. The study team will deposit de-identified quantitative and qualitative data along with study instruments into an open-access repository following University of Bradford and ethics committee guidelines after the study ends.

## Abstract

**Introduction:** The Southeastern region of Nigeria faces a severe public health challenge from hypertension due to its high prevalence. Psychological factors such as stress, anxiety, depression, coping mechanisms, and social support play essential roles in hypertension development and treatment, yet there has been limited scholarly attention to these factors in Nigerian and international qualitative research. This study addresses this evidence gap by investigating the psychological factors that influence hypertension development in adults living in Southeastern Nigeria.

**Materials and methods:** The sequential explanatory mixed methods study includes validated psychometric instruments assessing perceived stress, anxiety, depression, coping mechanisms, and social support. Regression analyses, correlation tests, and mediation/moderation models will be used to examine relationships among these variables. The research team will collect qualitative data through semi-structured interviews accessible to participants either online or in person. Thematic analysis following Braun and Clarke’s six-phase framework will be employed to explore participants’ lived experiences of stress, coping, and hypertension management.

**Ethics and dissemination:** Ethical approval was obtained from the University of Bradford Research Ethics Committee on 7^th^ August 2025. The research follows all procedures based on the Declaration of Helsinki and institutional ethical guidelines. Findings will be disseminated at academic conferences and published in peer-reviewed journals and stakeholder meetings held in both the UK and Nigeria. The research aims to generate evidence to support the development of comprehensive psychosocial care plans that address both physical and mental aspects of hypertension treatment in areas with limited resources.

## Introduction

The World Health Organization [1] reports that non-communicable diseases (NCDs), which include hypertension and mental health conditions like depression and anxiety, result in 74% of global deaths. Hypertension, popularly known as high blood pressure (BP), represents a significant worldwide public health emergency given that it stands as the leading cardiovascular disease, particularly in low- and middle-income countries (LMICs) [2]. The prevalence of hypertension in Nigeria has shown an alarming rise from 8.2% in 1990 to 32.5% in 2020 [3]. Recent meta-analytic research indicates approximately 57% of people aged 50 and above are currently living with the condition [4].

Hypertension is the most prevalent cardiovascular disease in Nigeria and leads to major emergency hospital admissions [3]. However, the increasing prevalence of hypertension does not translate to better awareness, treatment or control rates compared to high-income countries [5, 6]. Looking at BP measurements, sub-Saharan Africa, including Nigeria, consistently reports higher BP levels than European and North American populations, indicating a pressing need for enhanced approaches to hypertension care [7–9].

Current hypertension management approaches in Nigeria focus mainly on biological and socioeconomic aspects while largely neglecting psychological elements [10]. Yet a growing body of evidence shows that psychological factors, specifically chronic stress, anxiety, and depression play a major role in hypertension development and its progression [11, 12]. Further evidence reveals that psychological stress affects the autonomic control [13], hormone regulation [14], and medication adherence [15], which are fundamental pathways for managing hypertension. The incorporation of psychological screening alongside interventions into hypertension care practices represents an essential strategy to enhance treatment results for patient populations with high disease burden [16], namely, the Southeastern region of Nigeria which faces severe hypertension rates at 52.8% [9]. Southeastern Nigeria represents an essential research location for hypertension studies given the major disease burden and limited public understanding of the condition [17].

The global health community emphasises that cardiovascular care should include psychological health assessment by integrating psychosocial elements together with biomedical interventions [16]. However, even though a vast number of studies use quantitative methods to explore links between psychological distress and hypertension [e.g., 15, 18, 19], the literature lacks sufficient methodological depth because qualitative approaches are not adequately employed, thereby limiting how fully findings can inform clinical practice and service design [20]. Research depends mainly on quantitative measures and standardised scales to evaluate symptoms but fail to account for personal life experiences and social-cultural settings that influence psychological reactions to chronic illness [21].

The lack of psychological research in LMICs stands as a significant gap because these countries face structural, cultural, and economic barriers that neglect psychological factors affecting health [22, 23]. Several studies underscore the importance of addressing this deficit. Akinlua, Meakin [20] investigated hypertension beliefs among healthcare providers and patients in Nigeria and reported varied perspectives regarding its causes and treatments. Their findings demonstrate qualitative methods are needed to understand complex perspectives which help develop culturally appropriate interventions. Ukoha-Kalu, Adibe [24] found that patients and carers demonstrated minimal understanding of hypertension self-management and highlighted that family support alongside community and institutional backing played crucial roles in determining health behaviours which qualitative research methods can best analyse. Qualitative research serves as a fundamental tool to understand psychosocial elements [25] and cultural factors [26] that influence hypertension care in Nigeria. Such methods are essential to create targeted understanding and individualised treatments that adapt to local realities.

### Theoretical framework

#### Biopsychosocial model (BPS)

This research draws from the BPS which defines health as an interactive system between biological, psychological, and social factors [27]. The BPS model enables a complete study of hypertension which moves past physical measurements to study emotional states, coping skills and social relationships.

#### Health belief model (HBM)

As supporting frameworks, the HBM [28], explains how perceptions of illness severity and treatment barriers affect health behaviours [28].

#### Social support theories and stress-copping

The social support theory [29], and the transactional model of stress [30], further explains the importance of using the BPS structure. For example, the social support theory shows that supportive networks reduce stress [29] and improve treatment adherence [31], while the transactional model of stress and coping posits that people perceive and deal with stress as a dynamic process that affects hypertension management and treatment compliance [30].

#### Validated instruments further support these models

Consistent with the transactional model, coping style can moderate stress-related cardiovascular responses [32]. For example, studies have used the Coping Inventory for Stressful Situations scale [CISS-21 33] to assess how hypertensive patients cope when encountering stressful situations [34]. Further, studies grounded in the transactional model demonstrate that people who experience higher stress levels have a greater chance of developing hypertension [35, 36]. The Perceived Stress Scale (PSS-14) serves as a widely recognised tool for stress assessment [37] and has been used effectively in hypertensive patient groups and related risk populations [38].

According to the social support theory, people who receive social support tend to experience better BP responses when under stress [39] and lower hypertension risk [40], but its impact on hypertension control is context dependent [41]. Research indicates that social support affects both treatment compliance [42] and mental health outcomes [43]. Studies have used the Social Support Questionnaire [SSQ; 44] to measure perceived social support in hypertensive patients [39, 45].

Within the BPS psychological domain, research indicates that high levels of anxiety/depressive symptoms are associated with increased hypertension risk [12] and reduced treatment success through inadequate BP control [46] and non-adherence [47]. The Hospital Anxiety and Depression Scale (HADS) is a validated assessment tool for measuring anxiety and depression levels in patients with chronic diseases [48]. The HADS is widely used in these populations and shows connections to uncontrolled BP [10, 49].

Overall, the existing research demonstrates that psychological distress and psychosocial variables play essential roles in hypertension management, yet researchers and clinicians have not provided sufficient insight to these factors especially in LMICs.

### Study hypotheses

This research implements a mixed-methods methodology to study psychological elements affecting hypertension among the adult population in Southeastern Nigeria. The study is guided by the following hypotheses: (1) elevated psychological distress through stress, anxiety and depression will result in poorer hypertension outcomes, including elevated BP; (2) patients who feel more supported by their social networks will achieve better hypertension treatment outcomes; and (3) stress perception functions as a mediator between psychological distress, treatment adherence, with income levels and educational status influencing these relationships. The qualitative section of this study will enhance the quantitative results by examining how individuals experience psychological distress while coping with hypertension through the cultural lens of Southeastern Nigeria. This research will create context-specific knowledge that will develop patient-centred hypertension care systems for the Nigerian region and globally.

## Materials and methods

### Aims and objectives

Aim

To investigate psychological elements and social determinants that affect hypertension outcomes in Southeastern Nigeria with a goal of developing individualised and culturally informed approaches to hypertension management.

Objectives

1. To quantitatively establish specific connections between the independent variables—psychological distress (such as stress, anxiety, depression), social support, coping mechanisms, demographics, and the dependent variable—hypertension outcomes.
2. To qualitatively uncover deeper meanings and contextual understandings of individuals’ psychological experiences and their coping strategies in relation to hypertension.
3. To integrate research results into practical applications and policy development.

This protocol follows the STROBE (Strengthening the Reporting of Observational Studies in Epidemiology) guidelines for observational components and the COREQ (Consolidated Criteria for Reporting Qualitative Research) guidelines for the qualitative components of the study, as detailed in the accompanying Supporting Information files (S3 File: STROBE Checklist; S4 File: COREQ Checklist).

### Study setting and participants

The Southeastern Nigerian research location was chosen because (1) it shows one of the highest documented hypertension rates in Nigeria (e.g., ∼52.8% in community samples) [9]; (2) the population has low awareness and receives inadequate treatment and control [17, 50, 51]; (3) the area contains urban, semi-urban and rural settlements which support our research design and subgroup evaluation; (4) the existing partnerships in this region enable smooth participant recruitment and follow-up processes; and (5) the research addresses a regional evidence gap because, to our knowledge, no study has evaluated psychological factors affecting hypertension throughout all five Southeastern states.

Adults aged 18 years and older with confirmed hypertension diagnosis from a healthcare provider and/or currently taking medication prescribed to lower BP will make up the study population. The study does not require medical documentation for participant eligibility and does not use BP screening for enrolment purposes. Comorbid conditions and ethnicity do not restrict eligibility; these characteristics are recorded for analysis. The survey will ask participants to indicate how long they have had their hypertension diagnosis in four time periods: less than one year, one to five years, six to ten years, and more than ten years. Therefore, no minimum duration will be required for eligibility. All participants must live in one of the five states that make up the official Southeastern geopolitical zone of Nigeria which include Abia, Anambra, Ebonyi, Enugu, and Imo. Participants must be able to communicate in English as the interview and survey questions will be conducted in English. The study includes participants from urban, semi-rural, and rural areas to achieve demographic and socioeconomic diversity in the sample.

The study excludes participants who are under 18 years old, cannot communicate in English and live outside the Southeastern states, or have not reported hypertension through self-assessment (no previous diagnosis and no current antihypertensive medication).

### Study design

A sequential explanatory mixed methods research design will be implemented QUAN → QUAL [52]. An overview of the study design is presented in Fig 1. The quantitative phase follows a cross-sectional observational design, followed by qualitative data collection from a purposive subsample of survey respondents [53]. Integration occurs through linking different samples and uniting the findings during interpretation [54]. Qualitative strand interview participants will be chosen based on survey responses to achieve representation of essential quantitative groups [53]. For the interview guide, it follows a pre-determined structure based on transactional stress–coping, social support, and health belief models [28–30]. The quantitative section allows researchers to perform statistical evaluations of psychological factors which impact hypertension results (awareness, treatment, adherence, control) [11, 52]. The qualitative section gathers participant feedback about their stress management techniques [30], medication adherence, their understanding of hypertension and its treatment benefits/challenges [28], healthcare service interactions, and social support dynamics [29].

**Fig 1.** Overview of the sequential explanatory mixed-methods study design The study consists of three stages which are shown in the figure: (1) a quantitative cross-sectional survey was conducted to study the relationship between hypertension, psychological factors and behavioural patterns; (2) the study used semi-structured interviews to gather data about psychological experiences, coping mechanisms, and social support networks; (3) the integration phase combines quantitative and qualitative results to create culturally sensitive psychological approaches for hypertension care in Southeastern Nigeria.

Data collection began in November 2025 and is expected to be completed by July 2026.

### Sampling and recruitment

#### Sample size and power calculation

We used G*Power 3.1 to perform an a priori power analysis for linear multiple regression (test family: F tests; statistical test: *Linear multiple regression: Fixed model, R² deviation from zero*) [55]. The analysis used a medium effect size (f2 = 0.15), an alpha level of .05, and power of 0.80 [56]. The number of tested predictors was six: perceived stress, anxiety, depression, coping style, perceived social support, and sociodemographic factors. The G*Power analysis indicated a minimum of 98 participants to achieve the planned power, ensuring adequate precision for the proposed regression analysis while remaining practical and achievable.

To promote representativeness, recruitment will consider variation across the Southeastern Nigerian states, settlement types (urban, semi-urban, rural), and sex. Stratified sampling will be applied to achieve scientific rigour by providing equal representation of different sociodemographic groups, minimising sampling error, and enabling researchers to identify actual differences between population subgroups [57]. This approach also supports subgroup and interaction analyses involving sociodemographic covariates (e.g., whether stress affects men and women differently), while accounting for factors such as age, income, and education.

#### Sampling strategy

The quantitative phase will serve as the foundation for selecting participants through purposive sampling methods for the qualitative component (total sample = 30; sample selection includes 6 participants per state with 2 urban, 2 semi-urban, 2 rural areas, 1 male and 1 female participant in each area). This sampling approach aims to gather participants with diverse backgrounds including gender, age, income level, and educational status to make the study findings more transferable [53]. Qualitative interviews will continue until thematic saturation is achieved [58]. Research by Hennink, Kaiser [58] shows while code saturation can occur in 9-12 interviews, meaning saturation, which represents the point when no new insights appear usually requires 16 to 24 interviews specifically when working with diverse populations.

#### Recruitment procedures

The recruitment process will reach potential participants through healthcare facilities, community-based organisations, churches, and digital platforms that include social media and hypertension support groups in the southeastern region. The JISC-hosted survey platform will serve as the destination for all participants who receive recruitment materials including flyers, email invitations, and announcements. The JISC-hosted survey is mobile-optimised through smartphone-first design with large font sizes and basic navigation while using plain English language. Users can access the survey through a secure link which they can complete independently. Usability testing with 10 participants will be conducted before starting the main data collection process to achieve maximum completion rates.

### Data collection

#### Survey platform and instrument tool

The JISC platform hosts the quantitative data collection through a structured self-administered online survey in English only. The survey takes less than 30 minutes to complete and is divided into four sections about demographics and clinical aspects as well as lifestyle behaviours and psychological measurements. The full Participant Characteristics and Measures Questionnaire used in the quantitative survey is provided in S1 File.

#### Survey administration procedure

Using the JISC secure online link, all survey participants will receive an information sheet and provide consent before starting the assessment.

#### Participant characteristics and measures

The participant characteristics section contains 24 structured questions which are grouped into four sections titled Personal Information, Health Information, Lifestyle and Behavioural Factors, Geographic, and Cultural Context. All questions include checkboxes which allow respondents to choose their answers. Researchers developed the survey questions by combining proven instruments with standard national survey tools. The survey questions stem from the Nigeria Demographic and Health Survey (NDHS) [59], the WHO STEPwise Approach to NCD Risk Factor Surveillance [60] and the Nigeria Living Standards Survey (NLSS) [61]. These tools were chosen because they demonstrate cultural suitability alongside strong empirical evidence and wide application in health and behavioural studies across LMICs.

The BPS conceptual framework combined with the measurement framework adapted from NDHS, WHO STEPS, and NLSS allows researchers to gain detailed understanding about how psychological health and hypertension management relate to socio-demographic factors including behavioural and environmental elements in Southeastern Nigeria.

#### Personal information

This section collects essential information that provides foundational context for interpreting psychological and clinical outcomes. The assessment requests participants to share their information about age along with sex, marital status, educational background, employment status, income level, and residential area (urban, semi-urban or rural). These core indicators serve as essential elements for studying how social determinants of health affect hypertension-related behaviours together with psychological responses. Additional items assess participants’ living situation, occupational activity levels, and religious affiliations because these factors help explain gender dynamics, community traditions and health-seeking behaviours that are relevant in the Nigerian context [59–61]. Questions about healthcare service accessibility as well as past involvement in similar research projects to identify care challenges and research participation familiarity are also included.

#### Health information

The collected information focuses on participants’ treatment history related to hypertension. A formal high BP diagnosis serves as the initial eligibility criterion for study participation. Participants who qualify for the study must specify the time period since their hypertension diagnosis began (e.g., less than one year, 1–5 years). Additional questions evaluate family hypertension background, ongoing antihypertensive medication use, and existing physical or mental health conditions like diabetes and cardiovascular disease - adapted from the WHO STEPwise Approach to NCD Risk Factor Surveillance [60] alongside validated screening for anxiety and depression [e.g., HADS; 48]. The survey allows participants to select traditional remedies alongside standard pharmaceutical treatments including ACE inhibitors, beta-blockers, and calcium channel blockers [62, 63].

#### Lifestyle and behavioural factors

This section contains items which evaluate different health-related behaviours alongside psychological aspects connected to hypertension. Participants are required to state their exercise levels, their use of tobacco, alcohol as well as their dietary habits because these factors are confirmed risk behaviours for hypertension and cardiovascular disease [60]. The participants are asked to choose their perceived stress rating that best describes their experience [37] and identify their main coping mechanism when facing stressful situations among problem-focused, emotion-focused, and avoidant coping strategies [33]. These questions about coping mechanisms follow the transactional model of stress and coping [30] which evaluates coping responses through cognitive appraisal processes. The questionnaire employs a simplified format to make it easier for general population respondents to complete self-reports.

The final question in this section evaluates social support accessibility which demonstrates how interpersonal networks can buffer stress. The conceptual basis originates from social support theory [29]. For brevity and accessibility, the survey applies a single-item measure which aligns with the Social Support Questionnaire – Short Form constructs [64]. The survey items allow researchers to assess behavioural and psychosocial factors that are associated with hypertension related outcomes (e.g., BP, medication adherence) through both theoretical foundations and epidemiological evidence.

#### Geographical and cultural context

This section documents the state residence of participants within Southeastern Nigeria’s Abia, Anambra, Ebonyi, Enugu and Imo states which have recorded the nation’s highest hypertension rates [9]. It further includes voluntary religious affiliation questions which represent a typical demographic measurement in national surveys since cultural and spiritual beliefs strongly affect Nigerian health-related practices and psychological adaptation [59].

#### Psychological questionnaires

The psychological assessment component consists of four established/validated psychometric scales.

*The Perceived Stress Scale* [PSS-14; 37] assesses how participants perceive stress over the past month. It includes 14 items rated on a five-point Likert scale ranging from 0 (“never”) to 4 (“very often”). Higher total scores indicate greater perceived stress.

*The Hospital Anxiety and Depression Scale* [HADS; 65] assess symptoms of anxiety and depression. The scale contains 14 items that are arranged into two subscales (anxiety and depression). The scores produced will either read as normal (0–7), borderline (8–10) or signalling a possible clinical condition (11–21).

*The Coping Inventory for Stressful Situations scale* [CISS-21; 33] assesses how participants prefer to cope when encountering stressful situations. This research will use the 21-item version of the original 48-item CISS. Each subscale contains seven items that measure task-oriented, emotion-oriented, and avoidance-oriented coping strategies. The scores are rated on a five-point Likert scale ranging from 1 (“not at all”) to 5 (“very much”) conveying the identification of dominant coping patterns. The CISS-21 short-form version demonstrates acceptable reliability and construct validity when used with different populations and it remains suitable for brief survey periods because of its 21-item structure [66, 67].

*The Social Support Questionnaire* [SSQ6; 64] assesses the availability and satisfaction with perceived social support. The six items prompt participants to indicate the number of people they can turn to in specific situations and to rate their satisfaction on a six-point Likert scale, ranging from 1 (“very dissatisfied”) to 6 (“very satisfied”), with higher scores reflecting greater perceived support and satisfaction with that support network.

#### Qualitative data collection (interviews)

A purposive subsample of 30 survey participants who opt in to the qualitative component will be invited to take part in semi-structured, in-depth interviews. Interviews will be conducted either virtually through Microsoft Teams or in person at a hospital in Enugu and will last between 30 minutes and one hour. The researcher will use Microsoft Teams audio recording functionality to conduct online interviews after participants give their consent for recording. Next, the automatic transcript output from Teams will be examined and the necessary accuracy corrections will be performed. The researcher will use an encrypted digital audio recorder to record face-to-face interviews, while the host university stores all audio files and transcripts through its encrypted Microsoft OneDrive system which protects data during transmission and storage with password protection and limited access to the research team. The transcription process includes removal of names and contact information, and substitution with pseudonyms and Unique Participant Identification system (UPIDs) to achieve anonymisation. The research team will delete audio files after verifying the transcripts during a thirty-day period but will maintain anonymised transcripts for ten years according to the University of Bradford data retention schedule. Repeat interviews will not be conducted; each participant will complete one semi-structured interview. Transcripts will not be returned to participants for comment or correction, as member checking will occur through interview summaries rather than full transcripts.

The interview guide aims to investigate the psychological experiences of people with hypertension by examining their emotional responses to diagnosis and their stressors, anxiety, low mood, coping strategies, and social support networks. Cultural beliefs, socioeconomic limitations, and environmental elements will also be investigated to examine their effects on health practices and treatment compliance. The qualitative interview guide was pilot tested prior to full data collection to ensure clarity, cultural relevance, and appropriate flow of questions.

Participants will reflect upon personal experiences where psychological distress affected their BP management and treatment adherence. The qualitative questions in this study draw conceptually from the transactional model of stress and coping by Lazarus and Folkman [30], the SSQ [64], and the Perceived Stress Scale [37]. To ensure contextual relevance, the cultural and socioeconomic aspects are informed by the Nigeria Demographic and Health Survey [59], WHO STEPS framework [60], and research conducted in sub-Saharan African settings [9, 68]. The complete semi-structured interview guide is provided in S2 File.

#### Pilot study and study rigour

The research team conducted a pilot study before beginning full data collection to ensure survey instrument clarity, cultural relevance, and usability. The JISC-hosted questionnaire received responses from 10 hypertensive adults (two from each state). They provided their opinions about the survey’s language clarity, question understanding and time needed to complete the assessment. The research team also conducted cognitive debriefing to evaluate participant comprehension of the PSS-14, HADS, CISS-21 and SSQ-6 psychological scales. The pilot feedback led to small modifications in question wording which made questions easier to read and produced more response accuracy. The pilot test confirmed that participants were able to complete the survey between 25 and 30 minutes which validated both the study’s practicality and the survey’s appropriateness for the main research.

#### Qualitative rigour and trustworthiness

As the interviewer, AREO brings specific professional and academic characteristics that may shape the qualitative inquiry. She is a doctoral researcher in psychology with prior qualitative interviewing experience and an academic interest in the psychological dimensions of hypertension. She recognises that her background may influence how questions are framed and how participant narratives are interpreted. To minimise potential bias, she will engage in reflexive journalling, maintain awareness of assumptions, and participate in supervisory reflections throughout data collection and analysis. The research establishes trustworthiness criteria from Lincoln and Guba [69] to achieve quality and methodological rigour in its qualitative component. Credibility will be strengthened through iterative member checking, where participants may review summaries of their interview content for accuracy. The study will support transferability through complete descriptions of participant demographics and the Southeastern Nigerian setting which enable readers to determine how well the findings apply to their own research populations. Coding choices, theme development, and analytic procedures will be tracked through NVivo software to create an audit trail. Finally, confirmability will be ensured through reflexive journalling and supervision audit checks which will help reduce personal bias and support clear interpretation throughout the process.

#### Data management and confidentiality

All research data will be stored securely on an encrypted University of Bradford system, including password-protected Microsoft OneDrive, with access restricted to the research team. Personally identifiable information (e.g., names, email addresses, phone numbers) will be stored separately from research data and linked only through UPIDs. Survey data will be anonymised at the point of collection, while interview data will be anonymised during transcription to ensure that no identifying details are retained in the final dataset. The data cleaning process for the quantitative part will perform range and consistency checks, detect missing values, eliminate duplicate entries, and convert categorical variables to suitable formats for statistical analysis through dummy coding for regression models. The research team will create a complete data codebook to track all variable labels, their corresponding coding systems, and psychometric instrument scoring methods. For the qualitative component, NVivo software will be applied for systematic coding, and an audit trail will track coding decisions, revisions, and theme development history to maintain dependability and confirmability.

In accordance with the University of Bradford data protection policy, anonymised research data will be retained securely for 10 years. Personal identifiers will be deleted once the study and any necessary follow-up are completed, ensuring confidentiality and compliance with GDPR and institutional guidelines.

### Data analysis

#### Quantitative data analysis

Quantitative data will be analysed using IBM SPSS Statistics Version 29 [70]. Descriptive statistics, including means, standard deviations (SD), and frequencies, will be computed to summarise demographic characteristics and psychological variables. Pearson’s correlation coefficient (r) will be used to assess linear associations among continuous variables, provided assumptions of normality and linearity are met. Multiple linear regression analyses will be conducted to identify psychological predictors of hypertension-related outcomes, including perceived stress, medication adherence, and BP control. The evaluation of continuous outcome differences between groups (PSS-14, HADS-A and HADS-D scores, CISS-21 coping style scores, and SSQ-6 social support scores) will use independent-samples *t*-tests for two-level factors including sex (male vs female), treatment status (on treatment vs not), and residential setting when categorised into urban and rural categories. The analysis will use one-way ANOVA for factors with more than 3 levels, e.g., for residential setting, age group, state, education level, and income quintile.

#### Mediation and moderation analysis

The research will investigate social support as a mediator between psychological distress and treatment adherence through mediation analysis. The analysis of demographic moderation effects through regression uses interaction terms while ANOVA and regression require dummy coding for multiple category variables and *t*-tests use dichotomisation when theory supports it. The research will establish *p* < .05 (two-tailed) as its statistical significance threshold and report effect sizes with 95% confidence intervals to assess both clinical and practical significance.

#### Qualitative data analysis

We will analyse qualitative data using Braun and Clarke [71] six-stage thematic analysis method. The research process starts with (1) familiarisation, which involves multiple transcript readings for notetaking of initial ideas; (2) the NVivo software [72] will generate the initial set of codes by analysing the complete dataset to find essential elements which describe psychological and social experiences, and the primary researcher will conduct all initial coding, with coding decisions reviewed during supervisory meetings to enhance dependability; (3) codes will then be collated into potential themes by grouping patterns of meaning, and a coding tree will be developed iteratively as codes are organised into categories and preliminary themes; (4) themes will be reviewed and refined by checking coherence with coded extracts and across the entire dataset; (5) the research team will select vital themes which represent their core relationships to the research questions and then assign names to them; and (6) a complete narrative report which unites interpretive findings with supporting quotations from the data. Field notes will be made during and immediately after each interview to capture contextual details, non-verbal observations, and initial analytic reflections.

Themes will be developed using a combination of deductive (theory-driven) codes based on the biopsychosocial and stress-coping frameworks, and inductive (data-driven) insights emerging directly from participant experiences. The coding framework will undergo continuous development through supervisor meetings and audit trail analysis to achieve reliable interpretation results.

#### Triangulation of findings

The research design combines quantitative and qualitative data collection through triangulation to enhance validity and obtain context-specific insights [73].

## Ethics and dissemination

Ethical approval for this study was obtained from the University of Bradford Research Ethics Committee on 7 August 2025. The research follows the Declaration of Helsinki [74] and Beauchamp and Childress [75] ethical framework which supports respect for autonomy, provides benefits while avoiding harm, upholds justice and maintaining confidentiality.

All participants are required to give their full consent before starting the study. For the survey participants, they will show their consent by completing and sending the questionnaire after reviewing both the electronic participant information sheet and consent statement. The research team will get written or electronic consent from all interview participants before they start their participation. Participants will also be informed, through the Participant Information Sheet, that the interviewer is a doctoral researcher at the University of Bradford and that the study is being conducted purely for academic purposes. All participants will receive information about their ability to withdraw from the research at any time without facing any negative consequences.

The research data will be stored on University of Bradford systems which use encryption for protection under UK GDPR and institutional data protection policies. Personal identifiers will be stored separately from research data and linked only by coded identifiers. Quantitative data will be anonymised at the point of collection, and qualitative data will be anonymised during transcription to ensure confidentiality.

Anonymised data will be retained for 10 years in line with University of Bradford policy and securely deleted thereafter. The analysis will start after data cleaning and coding procedures are completed to achieve both quality and accuracy.

## Dissemination plan

This study forms part of a doctoral research project and the findings will be submitted as part of a PhD thesis at the University of Bradford. The research findings will be presented through academic journals, doctoral dissertation archives and international conferences which focus on psychology and public health.

Key findings and recommendations will also be shared with Nigerian health organisations, participating institutions, media, and community members through summary reports, policy briefs, and community meetings to enhance psychosocial and hypertension treatment in under-resourced areas.

## Data Availability

NA as study protocol
All data produced will be made available upon reasonable request to the authors

## Author Contributions

AREO: Conceptualisation, Methodology, Writing – Original draft. MC, MM and EV: Conceptualisation, Methodology, Supervision, Writing – Review & Editing.

## Supporting Information

**S1 File. Participant characteristics and measures questionnaire**

This file contains the full structured questionnaire administered through the JISC survey platform, including personal, health, behavioural, and contextual items.

**S2 File. Qualitative interview guide**

This file contains semi-structured interview questions that investigates psychological experiences, stressors, coping strategies and social support among adults with hypertension.

**S3 File. STROBE Checklist**

Completed STROBE checklist for observational study reporting.

**S4 File. COREQ Checklist**

Completed COREQ checklist for qualitative research reporting.

## Notes

### Competing Interest Statement

The authors have declared no competing interest.

### Author Declarations

Ethical approval was obtained from the University of Bradford Research Ethics Committee on 7th August 2025.

